# “Radiosensitivity Index” (“RSI”) is not fit to be used for dose-adjustments: a pan-cancer analysis

**DOI:** 10.1101/2021.08.13.21262017

**Authors:** Hitesh B. Mistry

**Affiliations:** Division of Pharmacy/Division of Cancer Sciences, University of Manchester, UK

## Abstract

**Purpose:** Re-analyze both the original preclinical and latest clinical pan-cancer open-source data-set to assess if the “Radiosensitivity Index”, “RSI” for short, explains enough of the outcome variance either preclinically or clinically to elucidate a dose-response empirically.

**Methods and Materials:** The original preclinical test-set data from the publication where “RSI” was derived was collected and re-analyzed by comparing the observed versus predicted survival fraction at 2Gy (SF2). In addition, the predictive capability of “RSI” was also compared to random sampling. Clinical data was collected from a recently published data-set that included “RSI” values, overall survival outcomes, radiotherapy dose and tumor site for 5 cancers, glioma, triple negative breast, endometrial, pancreatic and lung cancer. Cox proportional hazards model were used to assess: 1) is “RSI” better than random chance at ordering overall survival times; 2) does adjusting for “RSI” elucidate a dose-response and 3) does an interaction between “RSI” and dose exist.

**Results:** Preclinically “RSI” showed a negative correlation (Spearman’s rho = −0.61, p = 0.034) between observed and predicted SF2. Furthermore, 98 percent of random samples showed better correlation to SF2 than “RSI”. Clinically, the pooled concordance-index for “RSI” was 0.52 (standard error 0.2) i.e. it was found to be no better than random chance and so replicated the preclinical findings. Furthermore, a dose-response was not seen after adjusting for “RSI” (p=0.584) and no interaction between “RSI” and dose was found (p=0.147).

**Conclusions:** These results suggest that like the initial in-vitro analysis 12 years previously “RSI” is not a marker of radiotherapy sensitivity, should not be referred to as such and is also not fit to be used in any dose-adjustment algorithms.

## Introduction

The radiotherapy community has for decades been striving for an easy-to-use assay to measure the radiosensitivity of an individual patient’s tumor. Initially these approaches centered around developing short-term cultures (1) and lately the community has moved towards using omics data (2). The most prominent omics-based candidate biomarker over recent years is a marker termed the “Radiosensitivity Index”, “RSI” for short (3).

“RSI” plays a key role within GARD (Genomic Adjusted Radiation Dose), a formula that is being proposed to individualize a patients radiotherapy dose based on an individual’s cancers radio-sensitivity (4). GARD assumes that an interaction between “RSI” and radiotherapy dose exists. Furthermore, they implicitly assume that “RSI” captures enough of the variance in a response variable (e.g., overall survival [OS]) such that it elucidates a dose-response which can be used to tailor the radiotherapy dose to an individual. What do we mean by this statement?

Let’s return to the preclinical setting to expand on this point. Let’s assume we have 100 cell-lines, either patient-derived or from a cell-bank, and that each of these cell-lines produces a different dose-response within say an in-vitro radiosensitivity assay, see Figure 1. To generate the simulations, we used a linear-quadratic model and simply allowed alpha to vary and kept beta fixed for simplicity. If we had a perfect biomarker that fully explained the variance of the dose-response curves, once we account for that biomarker, all the dose-response curves would collapse onto one single dose-response curve. The biomarker would provide a link between the variance in response and dose that makes the dose-response look clearer. Thus, we could now use such a biomarker to dose-adjust with high degree of confidence of the resultant effect. If the biomarker was that good, we would only need a small variation in dose with a sufficient sample size to see part of the curve clearly. However, if the biomarker explains little of the variance, i.e., does no better than random chance, then a dose-response will never be seen regardless of the variation in dose.

**Figure 1:**
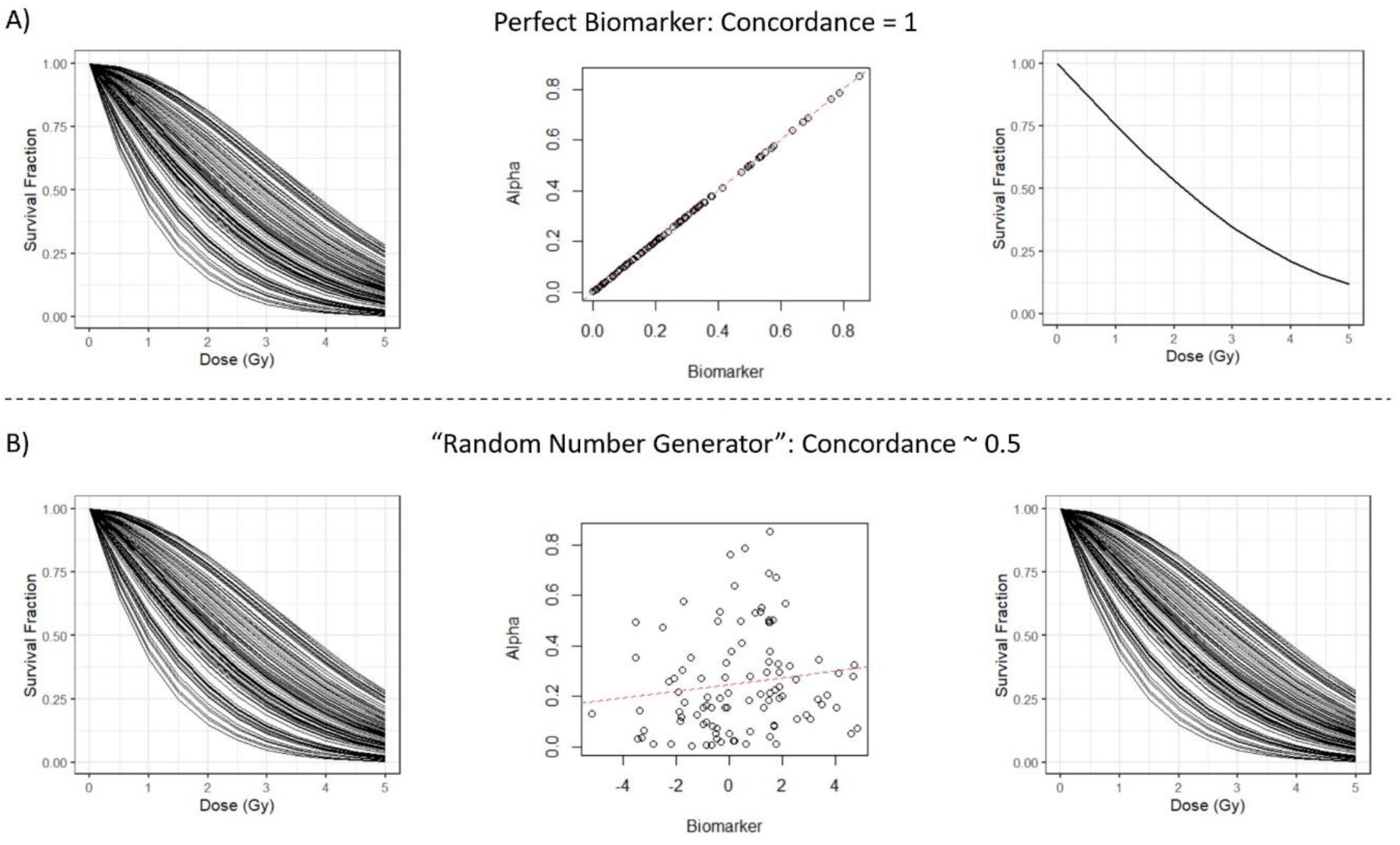
Plots showing how a biomarker that can perfectly predict each cell-lines alpha (concordance = 1) can collapse all the dose-response profiles to a single one (panel A) and a biomarker which is no better than random chance (concordance ∼0.5) cannot (panel B).

In reality the variance in a clinical end-point such as OS or any other time-to-event end-point will never be fully explained by any single biomarker. The question is whether enough of the variance can be explained such that a dose-response can be seen in the data. If we believe that we will never see perfect correlations (such as top-panel, Figure 1), how much of a correlation do we need to see given the dose-ranges used to see a dose-response? What other factors, which explain more of the variance, would we need to adjust for before we see a dose-response? It may be that a dose-response may never be seen. Note in pharmacology careful consideration has to be given for numerous sources of heterogeneity to have a chance of seeing a dose/exposure-response in efficacy and even then there is no guarantee (5, 6).

In this article we shall assess if “RSI” captures enough of the outcome variance such that it elucidates a dose-response but also assess if an interaction between “RSI” and dose exists. Neither of these analyses, which are key to assessing if “RSI” is suitable for use within a dose-adjustment algorithm, have ever been conducted.

## Methods

### Preclinical Analysis

The original test-set data used to develop “RSI” was taken from Eschrich et al. (3) The data was visualized by comparing the observed versus predicted SF2 and with Spearman’s rho reported with corresponding p-value. In addition, to assess how “RSI” compared to randomly guessing SF2 values a simple simulation study was performed as follows. 1000000 sets of uniformly random numbers, with each set the same size as the test-data-set, were generated. For each set Spearman’s rho was calculated comparing the random sample set to the observed SF2 from the test-set. These data were visualized using a histogram and the proportion of random samples that gave better agreement than “RSI” to the observed SF2 was reported.

### Clinical Analysis

“RSI” values, OS times (calculated from treatment initiation until death) and total radiation dose administered for patients who were treated with radiotherapy were taken from Scott et al. (7) (We refer the reader to that paper for further details on each cohort.) The triple negative breast cohort from Scott et al. (7) was not used as it only contained 9 events and such a low number of events can lead to large uncertainty in estimates of the survival probability without any predictors, see Figure 2. Any estimates from such a study are likely to be highly uncertain. Data on the melanoma study, reported in the original analysis (7), was not in the data-set provided at https://github.com/gsedor/GARD_Meta-Analysis at the time of writing and so that cohort was not considered either.

**Figure 2:**
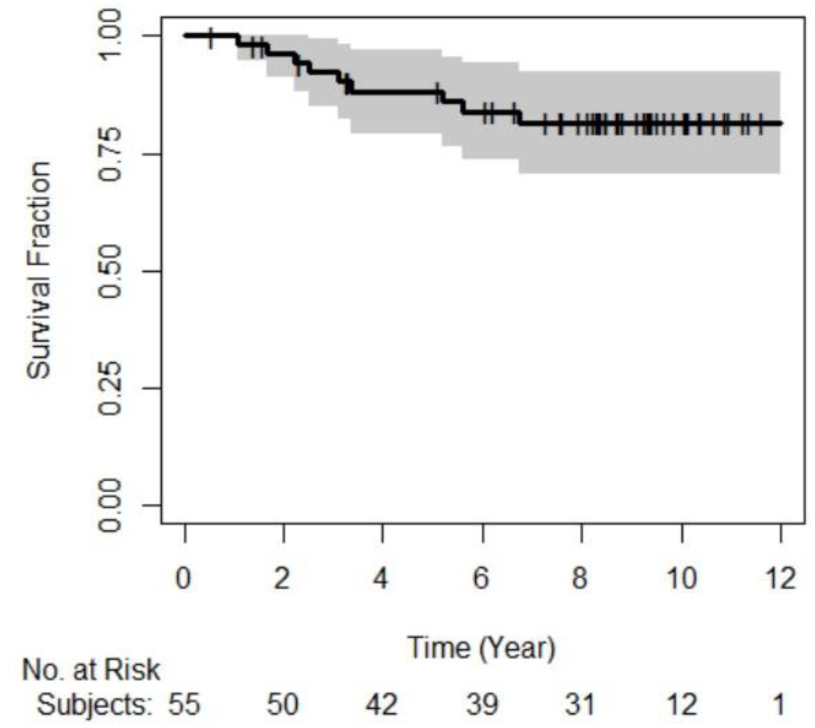
Kaplan-Meier plot of the triple-negative breast cohort. Vertical ticks represent patients who were right-censored and shed region is the 95% confidence interval.

For each tumor type, we first fitted a Cox proportional hazards (PH) model with “RSI” as the sole predictor and reported the concordance-index (c-index) with standard errors. Note a c-index value of 0.5 suggests that the predictor is no better than random chance whereas a value of 1 implies perfect concordance, recall Figure 1. Next, we considered a Cox PH model with both “RSI” and (total radiotherapy) dose as predictors. In this second model our interest was in whether dose becomes a predictor of OS after adjusting for “RSI”. If “RSI” explains enough of the variance in OS, then we expect to see good precision on the hazard ratio for dose. In this second analysis we shall report the hazard ratio of dose with 95 percent confidence intervals and p-value from the likelihood ratio-test assessing the fit between a model with “RSI” alone and a model with “RSI” + dose. Finally, for completeness, we assessed whether an interaction between “RSI” and dose exists by comparing a model with “RSI” + Dose to a model with “RSI” + Dose + RSI x Dose. (Note, GARD assumes an interaction exists here we assessed if an interaction term improved model likelihood rather than assuming it does.)

For all analyses, we considered each individual tumour site and a pooled analysis using tumour site as a stratification factor, i.e., the baseline hazard is different for each tumour site. That analysis assumes though there is no interaction between the stratification factor, tumour, and predictor, “RSI”, this may not be true. Therefore, we also assessed the no-interaction assumption by comparing a model with/without an interaction between tumor site and “RSI”, see Chapter 5 in Kleinbaum and Klein (8). Note that the no interaction assumption was not assessed in the original article by Scott et al. for any of their pooled analyses (7))

All analyses were performed in R v4.1.0.

Research data/code are available at: https://github.com/mcbi9hm2/PreclinicalRSI and https://github.com/mcbi9hm2/ClinicalRSICritique1

## Results

### Preclinical

Figure 3 shows the predicted versus observed SF2 values from the original test-set used to develop “RSI”. The plot shows that the performance of “RSI” is very poor, Spearman’s rho −0.61 (p=0.034) i.e. “RSI” appears to correlate negatively with its intended target, SF2.

**Figure 3:**
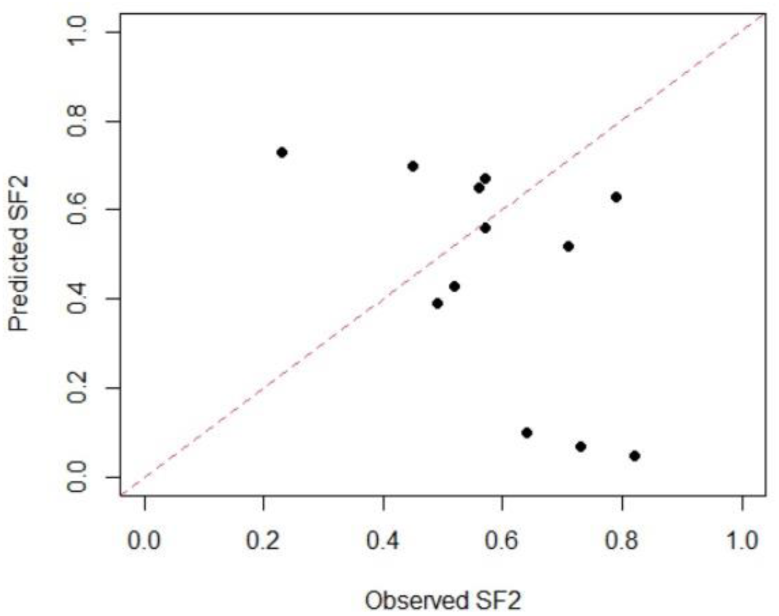
Plot showing the predicted versus observed SF2 of “RSI” using data from the test-set of the original publication. Dots are the cell-lines of the test-set and the red-dashed line the line of unity.

The results of the simulation exercise can be seen in Figure 4. The plot highlights that 98 percent of random samples have a better positive correlation to SF2 in the test-set than “RSI”. Preclinically, it appears that guessing an SF2 value is better than using “RSI” in the original test-set.

**Figure 4:**
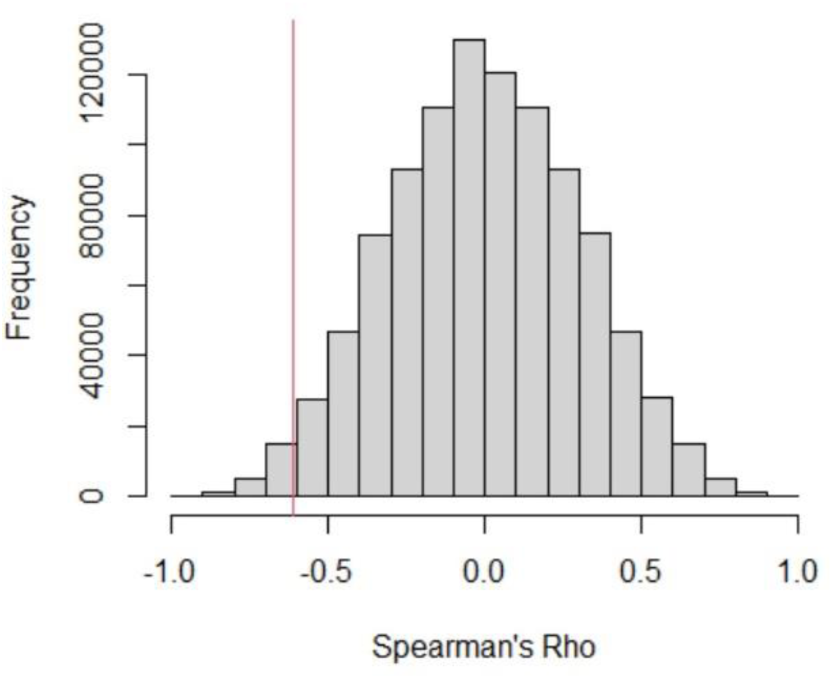
Plot showing the distribution of Spearman’s Rho values of random guesses for the test-set and the observed Spearman’s Rho for “RSI” (vertical red-line). A value of 1 would be perfect prediction.

### Clinical

Table 1 shows that “RSI” as a univariable predictor shows very poor discrimination, with low c-index values across all cohorts and in the pooled analysis. The interaction between tumor site and “RSI” was not needed (p=0.455). “RSI” does not appear to be better than random chance in any cohort and so explains none of the survival variance, i.e., we are in the bottom panel of Figure 1. Upon inclusion of dose, we see that “RSI” has not explained sufficient variance in OS to elucidate a dose-response. This second result is not surprising given “RSI” failed to explain any of the survival variance in a univariable analysis.

**Table 1:** Concordance indices for “RSI” in a univariable analysis and results of the bivariable model of “RSI” + radiotherapy dose

|  | “RSI” – C-Index (S.E.) | HR (95% CI) | LRT p-value |
| --- | --- | --- | --- |
| Endometrial | 0.55 (0.05) | RSI : 5.38 (0.24-122)<br>Dose : 1.02 (0.92-1.14) | 0.671 |
| Glioma* | 0.51 (0.03) | RSI : 1.09 (0.34-3.52)<br>Dose : 0.98 (0.89-1.08) | 0.664 |
| Lung | 0.55 (0.04) | RSI : 13.9 (0.52-366)<br>Dose : 1.02 (0.97-1.06) | 0.538 |
| Pancreas | 0.55 (0.06) | RSI : 2.57 (0.02-271)<br>Dose : 1.05 (0.88-1.25) | 0.585 |
| Pooled | 0.52 (0.02) | RSI : 1.78 (0.65-4.87)<br>Dose : 1.01 (0.97-1.05) | 0.584 |

\* Glioma this was unadjusted for MGMT expression. MGMT expression gave a c-index of 0.59, MGMT+“RSI” took this to 0.60. Dose still did not come out as a predictor LRT p-value 0.815.
LRT – Likelihood ratio-test, HR – Hazard Ratio, S.E. – Standard Error, CI – confidence interval

Table 2 shows that no interaction between “RSI” and dose exists in any cohort. Again, this result is not surprising given that “RSI” is itself not a predictor.

**Table 2:** “RSI”/dose interaction results

|  | LRT p-value:<br>“RSI” + dose + “RSI” x dose<br>v<br>“RSI” + dose |
| --- | --- |
| Endometrial | 0.382 |
| Glioma* | 0.681 |
| Lung | 0.297 |
| Pancreas | 0.496 |
| Pooled | 0.145 |

\* Glioma this was unadjusted for MGMT expression. Upon adjustment for MGMT expression an interaction between dose and “RSI” was still not seen (LRT p-value 0.605).

For completeness, the distribution of total radiotherapy doses is shown in Figure 5. Had “RSI” explained any of the outcome variance or showed an interaction with dose then a discussion around whether there was sufficient variation in dose and what causes the dose variance across these cohorts could have been had. However, this is not an issue if “RSI” explains none of the outcome variance and shows no interaction with dose.

**Figure 5:**
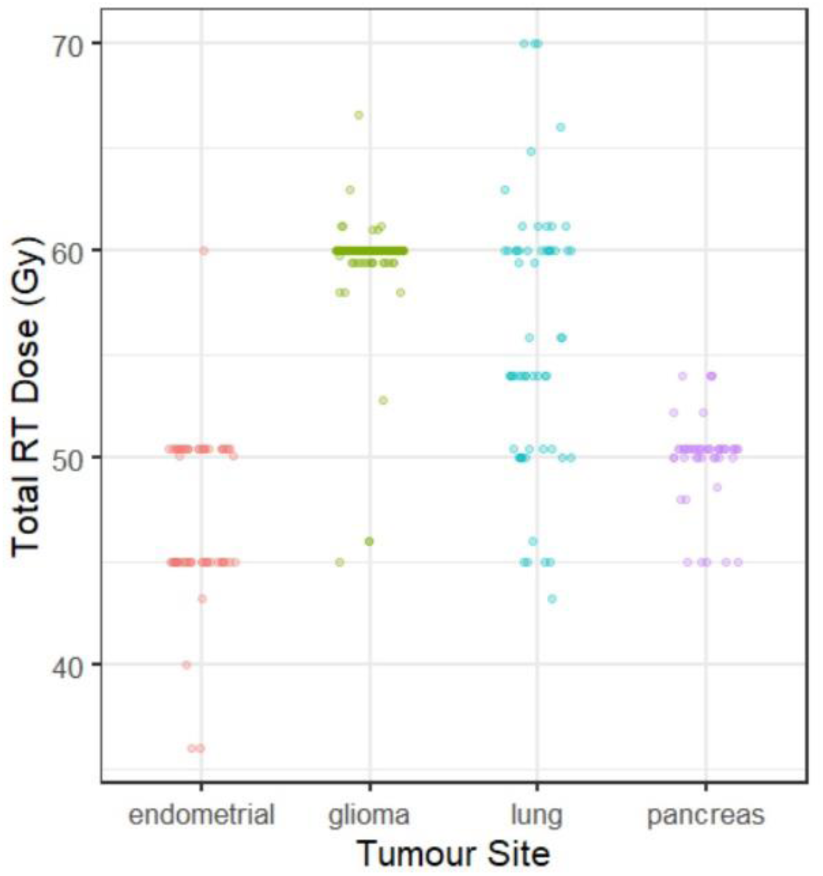
Distribution of total radiotherapy (RT) dose in the cohorts assessed here. Each dot is an individual patient.

## Discussion

Generating evidence that a biomarker(s) captures enough of the response variance (e.g., overall survival times) such that it can be used to make dose-adjustments is a hard task. In this article we have first highlighted what it means to capture enough of the response variance to elucidate a dose-response with a biomarker(s). We then re-assessed whether the preclinical and clinical evidence of a potential biomarker, termed “Radiosensitivity Index” (“RSI”), was able to elucidate a dose-response and also considered the less strict option of simply assessing if an interaction between “RSI” and dose existed.

Preclinically, we found “RSI” is no better than random chance at predicting the SF2 values within its original test-set. In fact, we found 98 percent of random samples showed a better correlation to SF2 than “RSI”. Therefore, “RSI”, does not explain enough of the outcome variance preclinically to elucidate a dose-response. (Note, the negative correlation between “RSI” predicted SF2 and the observed had been commented on previously but neither the developers of “RSI” or GARD have acted upon these comments (9).)

On the latest clinical data-set we found “RSI” explains none of the overall survival variance across numerous cancer types i.e. the results mimicked the preclinical findings of 12 years previous. Furthermore, since it did not explain any of the survival variance it also failed to elucidate a dose-response in any tumor type. Finally, we found there was no interaction between “RSI” and dose across any of the tumor types considered.

Both the preclinical and clinical findings have implications for the use of GARD as a dose-adjustment algorithm as we shall now discuss. Firstly, the preclinical results highlight that “RSI” is not an appropriate surrogate for an individual patient’s radiosensitivity value within GARD as it is unable to predict the SF2 values in a controlled in-vitro system. If a model cannot accurately predict radiosensitivity in a controlled preclinical environment, what chance does it have within in-vivo systems or clinically where numerous other variables are at play? The authors of “RSI” may argue that SF2 values are highly uncertain and thus the in-vitro data not reliable; in which case developing an algorithm on such data should not have been conducted in the first place and more robust preclinical data such as an in-vivo system should have been considered. Secondly, proposers of GARD assume that an interaction between “RSI” and dose exists but an analysis showing whether this interaction exists empirically was never done i.e. compared models with/without the interaction. Here we found that there is no interaction between “RSI” and dose. Thus, the assumption that there is an interaction between “RSI” and dose in GARD is not supported by the empirical evidence. These results highlight that GARD is not suitable to individualize patient radiotherapy doses in its current guise where “RSI” is used to predict a patients radiosensitivity.

It could be argued that one will never be able to personalize the radiotherapy dose for the following reasons. The variance in survival times is likely made up of numerous factors, ranging from type of combination therapy given with RT (which has its own predictors of response e.g., genomic factors, levels of drug in plasma etc.) but also standard clinical variables around patient fitness, follow-up care etc. and more. A breakdown how each variable contributes to survival times would be needed, together with how much can’t be explained, and then an assessment of how much of the variance due to radiotherapy can be explained by a candidate biomarker is needed. It may be more pertinent for the community to consider each disease in its own right, assess all available factors and decide whether they are currently explaining enough of the outcome variance to then pursue personalization of dose. It may be a personalization of both radiotherapy and systemic therapy doses.

In summary, this article highlights that “RSI” is not a marker of radiosensitivity, should not be referred to as such and is not fit to be used in dose-adjustment algorithms as is being proposed (7). Based on current evidence further research into “RSI” and dose-adjustment algorithms using “RSI” is not warranted and the community should move on.

## Data Availability

All data/code is available here: https://github.com/mcbi9hm2/ClinicalRSICritique1

## Notes

### Competing Interest Statement

The authors have declared no competing interest.

### Funding Statement

No funding was received.

### Author Declarations

This was a retrospective analysis of open-access data thus the analysis is exempt from needing IRB approaval.

